# Identifying vaccine-mechanism bias in mathematical models of vaccine impact: the case of tuberculosis

**DOI:** 10.1101/2022.10.18.22281211

**Authors:** M. Tovar, Y. Moreno, J. Sanz

## Abstract

In the development of vaccines against tuberculosis (TB), a number of factors represent burdensome difficulties for the design and interpretation of randomized control trials (RCTs) of vaccine efficacy. Among them, the complexity of the transmission chain of TB allows the co-existence of several routes to disease that can be observed within the populations from where vaccine efficacy trial participants are sampled. This makes it difficult to link trial-derived readouts of vaccine efficacy to specific vaccine mechanistic descriptions, since, intuitively, the same efficacy readouts may lean on the ability of a vaccine to arrest only some, but not all, the possible routes to disease. This increases uncertainty in evaluations of vaccine impact based on transmission models, since different vaccine descriptions of the same efficacy readout typically lead to different impact forecasts. In this work, we develop a Bayesian framework to evaluate the relative compatibility of different vaccine descriptions with the observations emanating from a randomized clinical trial (RCT) of vaccine efficacy, offering an unbiased framework to estimate vaccine impact even when the specific mechanisms of action of the given vaccine are not explicitly known. The type of RCTs considered here, conducted on IGRA+ individuals, emerged as a promising design architecture after the encouraging results reported for the vaccine M72/AS01_E_ clinical trial, which we use here as a case study.

**Authors summary:** Here, we focus on a problem that is pervasive in mathematical modeling of vaccines’ impact, consisting of the existence of a multiplicity of vaccine parametrizations that are compatible with the result of a given clinical trial of vaccine efficacy. However, focusing on tuberculosis vaccines, we find that it is possible to use computational simulations and Bayesian statistics to assign these models with posterior probabilities measuring their relative compatibility with the results of a real clinical trial under analysis. The framework presented unlocks the production of unbiased, mechanism-agnostic impact forecasts for vaccines against tuberculosis, and can be extended to the study of vaccines against other communicable diseases with a complex infectious cycle.

## Introduction

Despite the decay in TB incidence and mortality achieved worldwide since 1990 [1], its yearly rate of reduction is arguably too slow to meet the goal settled by the World Health Organization (WHO) in the End-TB strategy, which consists of completing a reduction of TB incidence and mortality rates by 90% and 95%, respectively, between 2015 and 2035 [2]. Instead, 2020 witnessed, for the first time in decades, an increase in global TB burden levels with respect to previous years, and that year the WHO estimated that TB was the cause of death of more than 1.5 million people worldwide, combining HIV negative and positive cases [3]. The cause of this increase was the irruption of the COVID-19 pandemics, which threatens, in countries like India, to raise the TB death toll back to even higher levels in the next few years [4]. This issue, as well as the ever-increasing rates of emergence of drug resistance [5], evidence the need of new epidemiological interventions and tools against TB; paradigmatically a new and better vaccine than the current bacillus Calmette-Guerin (BCG) [6], whose efficacy against the more transmissible respiratory forms of the disease in young adults is disputed [7].

Vaccine testing in TB is especially difficult due to a number of factors. They include the slowness of the contagion dynamics that forces vaccine developers to consider studies involving larger numbers of participants during longer follow-up periods than for other diseases [8-10], as well as the difficulty in defining trial endpoints for a disease where infection status can only be ascertained indirectly and immunological correlates of protection remain elusive [11]. This makes the design of RCTs for TB vaccines an extremely challenging and expensive task, in spite of which, nowadays, several preventive vaccine candidates against TB are being tested in human clinical trials [6,12]. Some candidates have completed phases 1, 2 and 2b of their development, and are about to enter into phase 3 to test their efficacy at providing prevention of infection (PoI) and/or prevention of TB disease (PoD) in large cohorts of thousands of participants recruited in high burden settings. In this context, the first phase 2-2b RCTs to be completed for new preventive vaccines against TB were those of the candidates MVA85A [8], M72/AS01_E_ [9,10] and also H4:IC31 [13], which were compared to a revaccination protocol with BCG. Remarkably, these three candidate vaccines, which collected disparate efficacy readouts, were tested within trials of noticeable diverse designs in several key characteristics such as geographical distribution, participants’ age or IGRA status, as detailed in table 1.

**Table 1.**
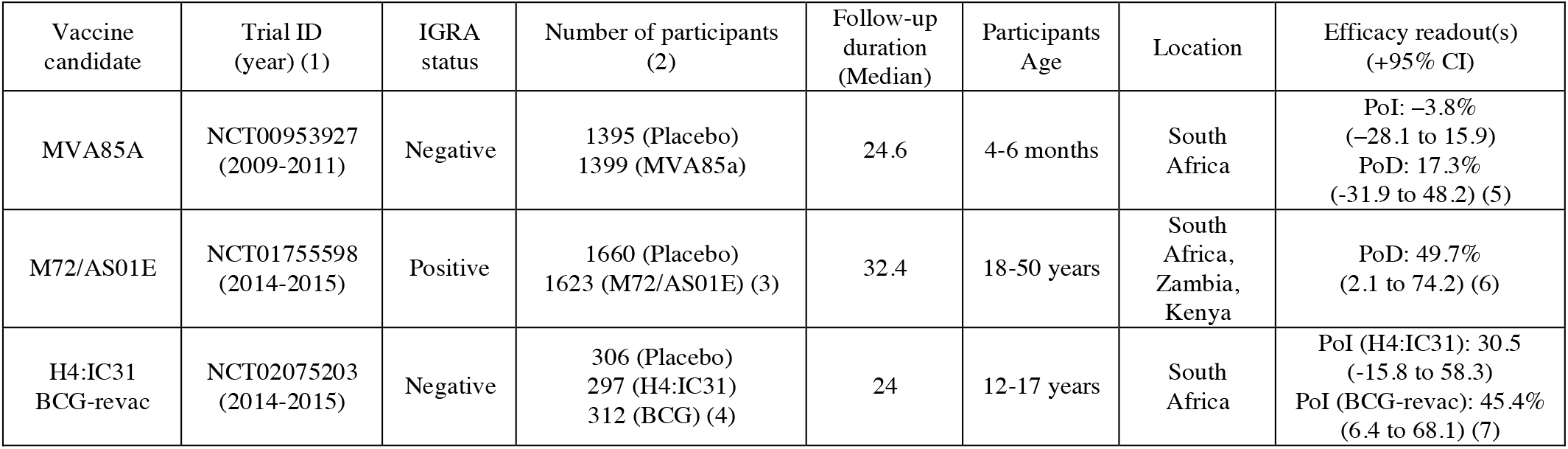
Phases 2/2b clinical trials for new TB vaccines. 1. Years of the recruitment phase. 2. Individuals included in efficacy analyses. 3. Individuals in according-to-protocol efficacy cohorts. 4. Individuals in per-protocol analyses. 5. PoD efficacy corresponds to endpoint definition 1, as described in [8]. 6. As reported in the final analysis [10], for the according-to-protocol efficacy cohort, first definition. 7. Efficacy against sustained QFT conversion (QFT conversion without reversion within 6 months).

The comparison summarized in table 1, involving just the first three pioneer phase 2/2b efficacy trials that have been completed so far, suggests, given the diversity of their designs, that the question of what is an optimal strategy for testing a preventive vaccine against TB at these stages of vaccine development lacks a unique answer, and that, as the rest of the vaccine candidates progresses through the development pipeline, the field will witness a higher number of trial designs being explored, as anticipated in [14]. This multiplicity of trial designs, along with the paucity of resources to allocate for evaluating novel TB vaccine candidates at a global scale [15], makes it absolutely critical to ensure that vaccines with different target product profiles, and, or estimated from trials of different characteristics can be timely compared in their expected ability to halt the global epidemics of TB.

One of the reasons why such a task is difficult is the fact that the PoI or PoD efficacy readouts obtained from an RCT do not offer an unequivocal characterization of a TB vaccine, since the same risk-reduction readouts observed in a trial can be mapped onto different mechanisms of action in different vaccine candidates [14]. This is extremely important because some of these compatible mechanisms are impossible to distinguish just by interpreting the trial’s results using standard methodologies, and yet, they appear associated to significantly different impacts, as foreseen by transmission models, if applied in simulated vaccination campaigns [16]. Yet, a majority of the models in the current literature on TB vaccine modeling base their forecasts on assuming that efficacy readouts unequivocally map onto specific combinations of action mechanisms without providing a plausible justification of this important modeling choices [17].

In this work, we propose a Bayesian modeling approach in order to relax such kind of assumptions. In our framework, we define a family of possible compartmental vaccine models characterized by different vaccine mechanisms from each of which we can estimate the likelihood associated to a particular RCT outcome. Using those likelihoods combined with uniform, non-informative priors for each of the possible models in the family, we can estimate the posterior probabilities of each model, providing in this way a means to evaluate the compatibility of each of the possible models with the RCT outcomes observed. Finally, we use these Bayesian posteriors as natural weights for each model’s impact forecasts, which -at least within the breath of the family of models considered-do not depend on mechanistic assumptions anymore.

To illustrate our approach, we analyze the case of the multi-centric clinical trial of the candidate vaccine M72/AS01_E_, conducted on IGRA-positive individuals from settings in three different high-burden countries: Kenya, Zambia and South Africa, which led to a promising PoD readout of *VE*_*dis*_ = 49.7% (95% *C. I*. 2.1 − 74.2). Applying our Bayesian formalism, we show that not all possible vaccine mechanisms that a priori could be included in All-or-Nothing compartmental models -arguably the most widely used type of vaccine models used in the modeling literature [17-19]-are equally backed-up by this specific trial readout. Also, we report that not all of them lead to comparable impact forecasts, which highlights the need of avoiding arbitrary choices in vaccine descriptions.

## Results

In a trial such as the one carried out for the vaccine M72/AS01_E_, conducted among TB-, IGRA+ individuals without a past of active TB, the episodes of incident TB to be observed during the study can be divided into three different groups, or routes to disease. First, some of the individuals whose IGRA conversion had occurred relatively recently will be expected to progress to primary TB during the first 12-24 months after exposure to the pathogen, which will typically overlap with the follow up period. Second, enrolled individuals whose IGRA+ status is associated with a latent TB infection (LTBI, linked to an exposure occurred, typically, >2 years before the beginning of the study) would be at a much lower risk of experimenting endogenous reactivation during the trial, although such events are still possible. Third, enrolled individuals may undergo primary TB followed upon re-exposure to the pathogen during the study. These three possible routes to active TB, sketched in the compartmental model diagram in figure 1A, are classically referred to as the “three risks model” [20], a frame coined by Vinnicky and Fine in 1997 [21]. In figure 1A we distinguish each of them according to one of the most commonly assumed model structures found in TB modelling literature [22], where LTBI individuals are split in fast (F) vs slow progressors (L), which we have chosen to describe the transmission dynamics of the placebo arms considered in this study.

**Figure 1.**
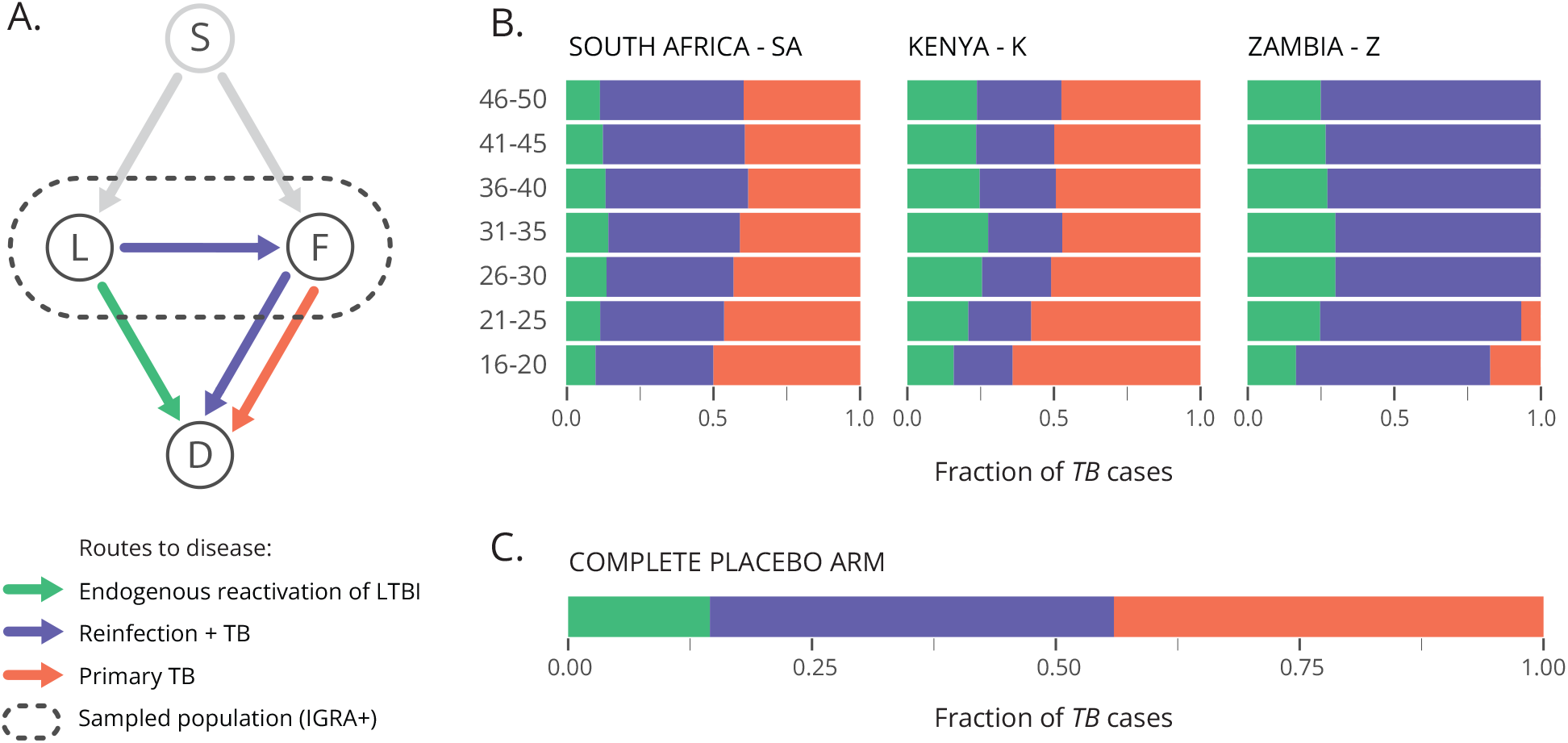
A priori characterization of the three routes to disease in the placebo arm of a Phase 2b clinical trial conducted on IGRA+ participants. (A). Compartmental model used to describe TB dynamics in the placebo arm of a trial conducted on IGRA+ individuals without past or present evidence of active TB. According to this model, trial participants can be divided in fast (F) vs. slow (L) progressors, each of which show different risks of progression to disease (D) per unit time that can be further divided into three routes to disease. (B) Fraction of incident TB cases that can be expected to correspond to each of the routes to disease in simulated trials conducted on each country and age-group. (C) Weighting the contributions estimated in panel B, according to the age and country-wise distributions of participants in the M72/AS01_E_ trial [9], we obtain an overall estimate of the relative contribution of each route to disease to the total incidence observed in the global placebo arm of the trial.

From this model description, our first goal is to implement computational simulations to estimate the relative weight of each route to disease in the placebo arm incidence of a clinical trial such as the M72/AS01_E_ study. To implement such simulations, we need two main ingredients: the epidemiological parameters governing the transitions in figure 1A, and the expected initial prevalence of fast (F) vs. slow progressors (L) in the population sampled for the study. To gather estimates of these key parameters, we calibrate an exhaustive TB transmission model previously developed [23] in each of the countries involved in the study, from where we retrieve estimates of the epidemiological parameters that play a role in the minimal model of figure 1A, as well as the relative prevalence of fast (F) vs slow (S) individuals in the population corresponding to 2015, the year that the M72/AS01_E_ study took place (see Methods and Supplementary figure S1).

With those ingredients at hand, we perform *in-silico* trial simulations in each country and age-group, wherein participants’ fates are simulated stochastically, according to an implementation of the Gillespie algorithm that allows tracking the three routes to disease independently (see Methods and SI, table S1). In our case, we use the multi-centric trial of M72/AS01_E_ as a case study [9,10], which enrolled participants between 18 and 50 years old in three countries: South Africa, Kenya and Zambia (figure 1B). Finally, using these results along with the demographic profiles of participants reported in the M72/AS01_E_ trial [9], we can produce a global estimate of the contribution of each route to disease to the incidence observed in the placebo arm of the entire study in the M72/AS01_E_ trial, as an average of the results of the age groups and countries involved in the study weighted by their relative frequencies in the trial (figure 1C).

Now, within the framework of the “three risks model”, it is conceptually possible that vaccines may provide PoD by reducing only, or preferentially, the disease risk associated to some of these three routes to disease. The estimates of the relative share of total incidence that can be attributed to each route to disease gives us very useful information about what are the precise mechanisms that may be more interesting to target in a given population. However, the immunological components of host responses against *Mycobacterium tuberculosis* (*M*.*tb*., the causative agent of TB). that are involved in protecting against primary TB upon recent infection, endogenous reactivation, or re-infection are complex and neither homogeneous nor linear [24]; and could be boosted to different extents by a vaccine in a way that is extremely hard, if not impossible, to predict a priori.

To accommodate modelling decisions to this uncertainty we consider a set of vaccines that provide PoD by reducing each of the three individual risks, either alone, or combined (figure 2).

**Figure 2.**
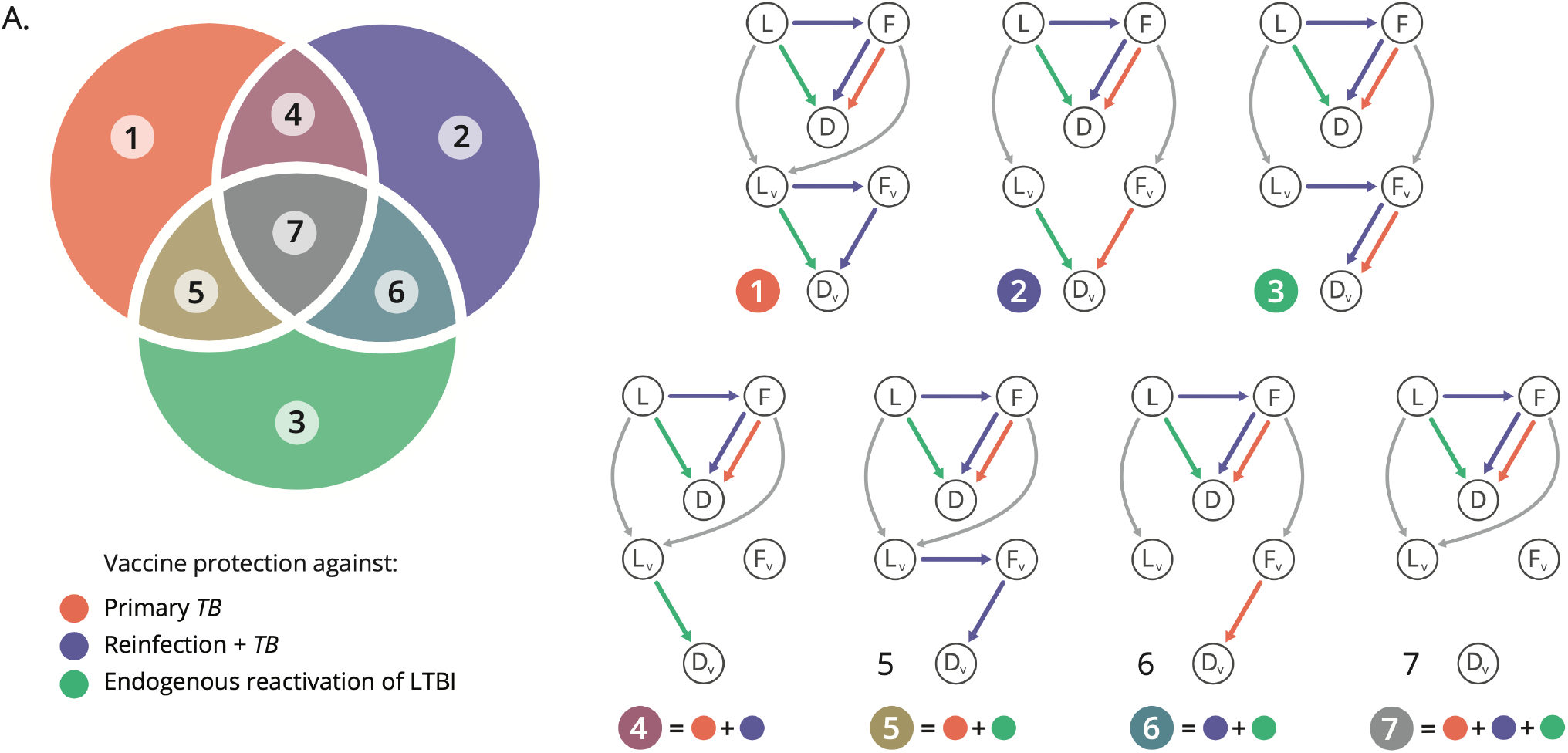
Compartmental models to accommodate the description of vaccines providing PoD by acting on specific routes to disease. (A). Venn diagram sketching the seven vaccine types contemplated in the study, as a function of the routes to disease they are assumed to protect against: primary TB (model 1), TB upon reinfection (model 2) or endogenous reactivation of LTBI (model 3). Combinations of these mechanisms yield models 4-7, which describe vaccines that are able to halt two (models 4,5 and 6), or all three routes to disease at once. Each region of the Venn diagram corresponds to a value of the discrete index *i* ∈ {1,2, …, 7}. (B) Compartmental descriptions of each vaccine type. In each of the seven models, a vaccine arm is included in parallel to the placebo arm, that defines the disease dynamics of the vaccinated individuals who are protected against developing disease through the corresponding routes.

This yields seven vaccine models, that can be denoted as *M*(*i, ε*), where we have each model being defined by the integer index *i* ∈ {1,2, …, 7}, determining the specific protection mechanism(s) present in the vaccine (figure 2A) and the continuous parameter *ε* ∈ [0,1], which captures the intrinsic efficacy, modelled as the fraction of individuals protected within an all-or-nothing modeling framework, considered identical for all the vaccine effects present in each case.

This way, while in models 1-3 only one of the three routes to TB is disrupted by the vaccine, models 4-7 incorporate several mechanisms simultaneously (figure 2). For instance, model 5 describes a vaccine that protects against primary TB and against LTBI endogenous reactivation at the same time. Therefore, the set of vaccine descriptions {*M*(*i, ε*)} will constitute a space of possible models, within which we will look after the one(s) whose assumptions are most compatible with a trial’s PoD readout of vaccine efficacy, that is, with the largest Bayesian posteriors, instead of blindly assuming that a vaccine acts through a given specific mechanism, or, for example, that it reduces all risks alike. To accomplish that task, we expand the Gillespie stochastic simulations mentioned before to include the simulation of vaccine arms each of them coherent with the seven types of vaccine models described above (figure 2B).

To describe vaccine behavior in the individuals assigned to the intervention cohort in each of the seven vaccine types, we assume a uniform non—informative prior on the efficacy parameter, and register the observed efficacy against disease *VE*_*dis*_ (that is, the PoD readout) that is associated to each trial simulation (see figure 3A). From these results, we can estimate the likelihood *P*(*VE*_*dis*_= 49.7% |*i, ε*) associated to each possible model defined by the combination of parameters {*i, ε*}. This likelihood can be interpreted as the probability that a trial conducted on a given simulated vaccine will lead to a PoD efficacy readout *VE*_*dis*_that is compatible with the observations made in the real trial: that is, in our example, *VE*_*dis*_= 49.7% [10].

**Figure 3.**
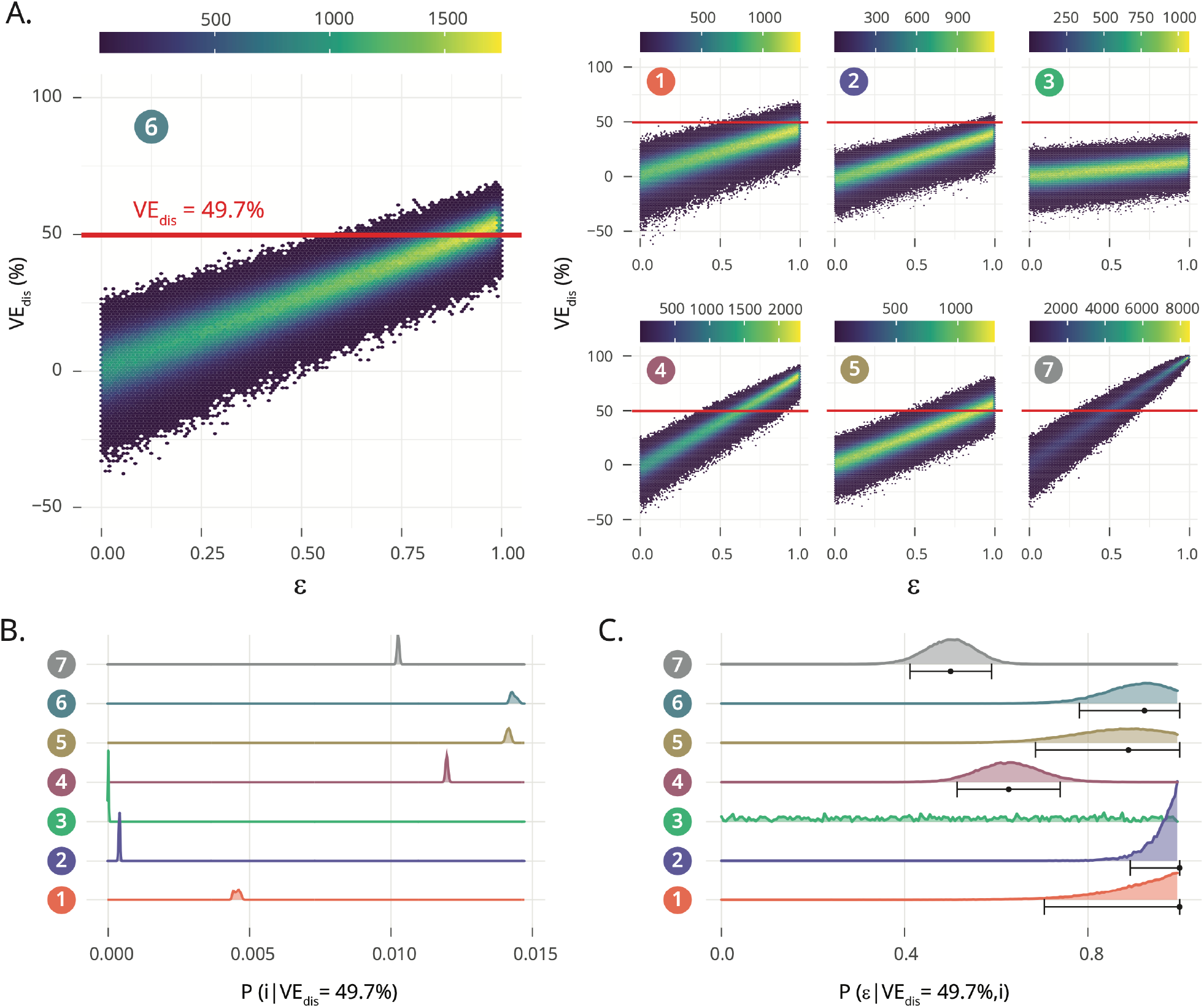
Bayesian analysis of possible modeling architectures underlying a trial-derived observation of vaccine efficacy. (A) Absolute frequency density distributions of efficacy values *VE*_*dis*_ obtained in sets of N = 2×10^6^ clinical trial simulations per model, uniformly distributed across the intrinsic vaccine efficacy parameter *ε* (efficacy resolution: 0.005, with 10.000 realizations for each value of *ε*). Red horizontal lines mark the PoD efficacy observed in the M72/AS01_E_ trial *VE*_*dis*_= 49.7%. (B) Marginal posteriors *P*(*i*|*VE*_*dis*_= 49.7%), capturing the relative compatibility of each model with respect to the efficacy observed in the M72AS01_E_ trial. (C) Distribution *P*(*ε*|*VE*_*dis*_= 49.7%, *i*) of the intrinsic vaccine efficacy parameter *ε* in each model type, given the observed efficacy *VE*_*dis*_= 49.7%, along with mean and 95% confidence intervals associated to them. For M3, the CI was omitted, for it spans the entire range *ε* ∈ [0,1], as the model fails systematically to produce simulation instances compatible with the observed *VE*_*dis*_= 49.7%.

Using this likelihood term, we can apply Bayes rule to define the posterior probability associated to each particular model, *P*(*i, ε*|*VE*_*dis*_= 49.7%), and derive a marginal posterior probability over the model type *P*(*i*|*VE*_*dis*_= 49.7%), by integrating *P*(*i, ε*|*VE*_*dis*_= 49.7%) over all possible intrinsic efficacy values *ε*. These marginal posteriors *P*(*i*|*VE*_*dis*_= 49.7%), represented in figure 3B, provide a mean to quantify the relative support in a given trial’s outcome for each one of the seven different vaccine descriptions provided. In our case, the observed PoD efficacy readout reported for the vaccine M72/AS01_E_ [10] appears more compatible with models 4, 5, or 6, each featuring a combination of several vaccine mechanisms, than with models where vaccine effects are associated to a unique mechanism of action. Interestingly, the model showing the greatest posterior probability is not the model where the vaccine is assumed to interrupt equally the three process, in spite of this model architecture being the one used in most recent modeling works aimed at producing impact estimates for TB vaccines similar to M72/AS01_E_ [25,26].

Our approach can also be used to estimate the intrinsic efficacy values *ε* that are most compatible with the observed PoD efficacy readout *VE*_*dis*_= 49.7% under each of the model type descriptions, this time applying the Bayes rule over each of the seven types of vaccine models independently to obtain the conditional posteriors *P*(*ε*|*VE*_*dis*_= 49.7%, *i*). The first momentum of these conditional posteriors corresponds to the expected values of the intrinsic efficacy parameter under each model type, that is ⟨*ε*⟩_*i*_, which is captured in figure 3C, along with its confidence intervals obtained from the conditional posterior distribution *P*(*ε*|*VE*_*dis*_= 49.7%, *i*) itself. These expected efficacy estimates illustrate a sensible feature of our model approach, namely, that a given PoD readout *VE*_*dis*_must be mapped to lower intrinsic efficacy *ε* values when the vaccine is able to halt progression to disease through all possible routes than when it acts on a subset of them.

Once we have described our Bayesian approach to inform vaccine characterization combining trials’ results with in silico simulations, we illustrate how it can be used to reduce arbitrariness from impact evaluations based on transmission models. A typical line of action to prospective impact evaluation of a vaccine consists of three steps: 1) implementing a transmission model accommodating a sensible vaccine description defined a priori. 2) Infer the vaccine parameter(s) conditional on the model structure that provide an optimal agreement with trials data; and 3) produce model-based forecasts of vaccine impact. A potential problem with this approach is, of course, that there exist many vaccine descriptions that can be adopted in the second step, and that they may lead to substantially different impact forecasts.

In order to illustrate this problem and quantify its importance, we capitalize on the same transmission model used above to infer epidemiological parameters and fractions of fast vs. slow progressors. This model [23], was later adapted to allow for the description of the effects of the introduction of new vaccines [16]. Here, we have further adapted the model to accommodate vaccine descriptions compatible with each of the seven models under analysis (see Methods & SI).

In figure 4A, we see the incidence reduction rate (IRR), evaluated in 2050, achieved by the introduction of a vaccine in 2025, on a vaccination campaign targeting teenagers (16-20 years old), under each of the seven types of models analyzed in this study in three high burden countries: India, Indonesia, and Ethiopia. Here, the intrinsic efficacy modelled in each case corresponds to the expected value ⟨*ε*⟩_*i*_ conditional to the model architecture and the vaccine trial PoD readout *VE*_*dis*_ observed in the trial. Importantly, we observe statistically significant differences in impact between models, as seen in figure 4B, where we illustrate the relative differences in IRR foreseen in each country by each vaccine model and the maximum a-posteriori model 6, describing a vaccine that protects against endogenous reactivation of LTBI and TB upon reinfection at once. These differences are statistically significant in 11 out of 18 cases (Bonferroni-adjusted p-values <0.05), and account for as much as 77.6% of the impact foreseen by model 6 in the most extreme case, (*IRR*(3, ⟨*ε*⟩_3_) vs *IRR*(6, ⟨*ε*⟩_6_) in Indonesia). These results evidence the importance of removing arbitrariness from modelling choices of vaccine descriptions.

**Figure 4.**
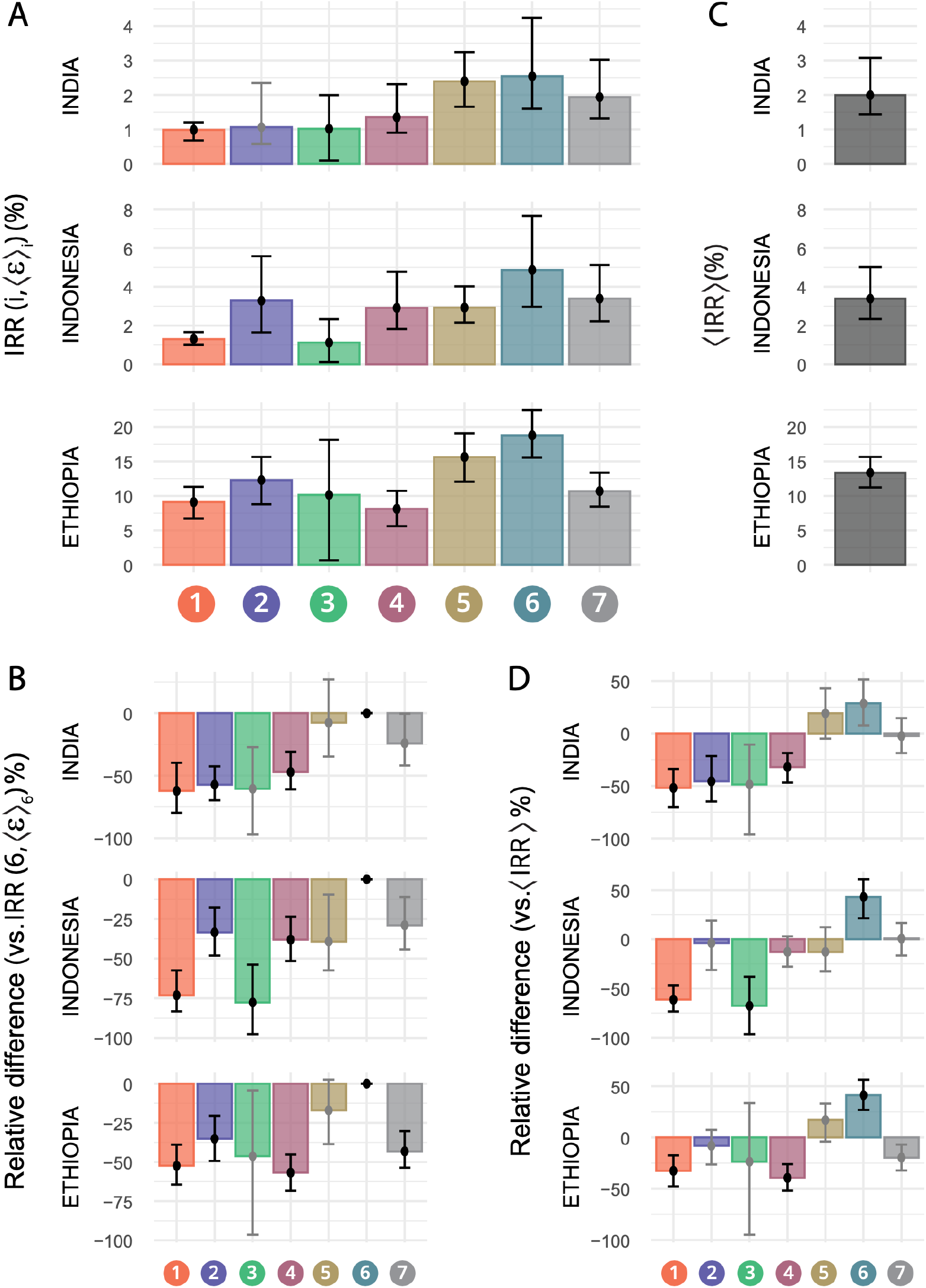
Impact forecasts variation across model structures vs. mechanism agnostic Bayesian estimates of impact. (A) Vaccine impact forecasts obtained through the comprehensive transmission model introduced in [16,23], when the vaccine is modelled according to the each of the seven descriptions here discussed. (B) Relative differences between the impacts foreseen by each model and the model with maximum Bayesian posteriors (model 6). (C) Combined, mechanism-agnostic ⟨*IRR*⟩ estimates for the same impacts, in the same countries, where each of the seven models contributes proportionally to its Bayesian posteriors. (D) Relative differences between ⟨*IRR*⟩ and impacts foreseen by each individual model. Black error-bars correspond to significant statistics (adjusted Bonferroni p<0.05, with N=63 tests).

This can be achieved, at least within the family of models under analysis, using our Bayesian approach. In short, we propose considering Bayesian estimates of expected vaccine impact ⟨*IRR*⟩ as a mean of the impacts foreseen by each type of vaccine *IRR*(*i*, ⟨*ε*⟩_*i*_), weighted by the marginal posteriors *P*(*i*|*VE*_*dis*_= 49.7%). The results of this exercise are presented in figure 4C for India, Indonesia and Ethiopia. In the impact forecasted in figure 4C, IRRs range from 13.35% in Ethiopia vs 1.99% in India, in line with results provided in other recent modelling studies for vaccines of comparable profiles, in comparable vaccination strategies [25,26]. As with comparisons across models, deviations of individual vaccine descriptions with respect to ⟨*IRR*⟩ range between +42.93% (*IRR*(6, ⟨*ε*⟩_6_) above ⟨*IRR*⟩ in Indonesia) and -96.30% (*IRR*(3, ⟨*ε*⟩_3_) below ⟨*IRR*⟩ in India), and are statistically significant in 9 over 21 occasions (Bonferroni-adjusted p-values <0.05, figure 4D). This highlights again the risk of adopting a priori a given dynamical structure for vaccine descriptions in transmission models, and the convenience of adopting a Bayesian approach on this problem as we propose here.

## Discussion

In a disease with a complex transmission chain, such as TB, vaccine mechanisms can be modelled in many different ways, some of which can be rendered compatible with clinical trial observations and yet produce divergent results when plugged into transmission models for their prospective evaluation. To solve this problem, we propose a method that combines *in-silico* simulations with actual trial results to quantify the relative compatibility of different vaccine descriptions with trial-derived observations. These model-to-data compatibility metrics are nothing but Bayesian posteriors which we use as weights to retrieve expected vaccine impact forecasts where models that are more compatible with trial observations contribute more than those in conflict with data. By doing this, we provide a method that helps circumventing the need to make arbitrary modelling decisions with respect to vaccine mechanisms, which may bias their quantitative conclusions.

The discussion addressed here is pertinent within the context of TB vaccines development, since vaccines activating certain immune pathways and responses may exert, simultaneously, different effects on the risk of developing TB associated to different routes to disease. This is because of the complexity of the successful host responses to TB, which are commonly grouped around two different strategies, termed resistance (the ability of the host to clear infection, typically early after pathogen entrance to the airways); and tolerance (the ability of the host to contain the pathogen indefinitely) [27]. While resistance typically depends on the existence of strong early innate responses, coordinated with the early participation of conventional CD4+ T cell responses, the importance of other immunological components for a successful resistant phenotype, such as unconventional tissue resident T cells, as well as NKT, NK and possibly B cells, is increasingly recognised [24]. However, while the strength of these responses is probably needed to permit early pathogen clearance, when the pathogen evades them and latent infection stablishes, host tolerance depends on their ability to modulate the strength of many of the very same response components required for guaranteeing resistance. CD1-restricted unconventional T cells constitute an illustrative example of this: while their ability to sense *M*.*tb*. cell wall lipids has been linked to the resistant phenotype [28], they also target mitochondrial associated lipids released by the host under stress conditions, which can therefore lead to tissue damage in the lungs and cavitation in the later phases of the infection cycle, ultimately leading to loss of tolerance, reactivation of TB, and transmission [29,30]. Considering the paramount complexity of the problem, when modeling vaccine effects, one should favor frameworks that minimize the amount of a priori assumptions needed about vaccine mechanistic behavior, such as the one here proposed.

As a case example of the potential of our approach, we analyzed the phase 2b efficacy trial of the promising vaccine candidate M72/AS01_E_, conducted on individuals previously exposed to the pathogen (IGRA+). Here, we produced weighted averages for the impact of this vaccine, to conclude that M72/AS01_E_ is expected to lead to an IRR of 13.35, 1.99 and 3.38%. by 2050, in Ethiopia, India and Indonesia, respectively, if applied on adolescents uninterruptedly, starting in 2025. These impacts are modest, implying that wider vaccination campaigns would be necessary to meet the End-TB strategy goals if the efficacy profile of this vaccine is consolidated in further, phase 3 studies and no better tool is at hand.

Using our method, we found that the vaccine model offering the highest posterior probabilities given the trial results is model 6, where vaccine PoD leans on protection against endogenous reactivation of LTBI and TB upon reinfection. This result is interesting, for it suggests that vaccine effects on halting on-going fast transition to TB on individuals infected not long before vaccination (the only mechanism absent from model 6) are not more important, or likely to be present in this particular vaccine with the data at hand, than the other two. This is a reassuring observation, especially given that the M72/AS01_E_ candidate includes the powerful liposomal adjuvant system AS0E1 [31] that was not supplied to participants in the placebo arm of the study. This feature of the trial design has the advantage of providing a comparison to a *clean* placebo arm, at the cost of not precluding the possibility that some of the protection observed may not be a consequence of the (adjuvated) vaccine efficacy, but a direct effect of the transient immune priming caused by the adjuvant itself. While ruling out this possibility is important, the analyses presented here are arguably insufficient to that end, since other models that include a vaccine effect on halting TB progression of recently infected individuals (e.g. models 4,5), show comparably high Bayesian posteriors (figure 3B). In this sense, the potential of our approach to disentangle specific vaccine mechanisms with better specificity than what is presented here could be further exploited if applied to the analysis of multi-centric RCT with divergent TB demographics and, unlike the example analyzed here, homogeneous representation across sites. Using our method to interpret, in parallel, RCT data compiled at epidemic settings with different expected distributions for incident TB cases across routes to disease would unlock interrogating for the correlation between those distributions and the Bayesian posteriors of the different vaccine models proposed.

The approach here introduced fosters important limitations. First, the implementation of the clinical trials simulations requires estimating a series of epidemiological parameters a priori, including the fraction of individuals in the fast vs slow progression reservoirs, rates of re-infection, and fast progression to disease; all conditioned (at least) by age stratum and epidemic setting; in order to combine them, at a later stage, to describe the global study population. This was done in this study by assuming unbiased representation of the age groups in each country in the study cohorts, and assuming that the overall epidemic risk in the countries of the trial were representative of the overall situation in each specific setting in the year of the study. Admittedly, exact age distributions of the participants (whenever available, and possibly complemented with further information about risk factors), and more relevant information on incidence levels at the specific settings could be used in order to refine quantitative conclusions. Similarly, IGRA+ clinical trial designers should include strategies to explicitly quantify the fraction of the participants who underwent recent vs remote IGRA conversion before the beginning of the trial, which would remove the need of estimating fast vs slow progression reservoir from transmission models. This could be done directly (i.e. by including an IGRA screening phase lasting circa one year before trial starts, where individuals who are initially testing negative are periodically re-evaluated to capture a fraction of recent IGRA conversions before the beginning of the study), or, perhaps more feasibly, by using biomarkers of time since IGRA-conversion, a promising possibility that is technically available, as recently demonstrated in [32]. Finally, it is important to highlight that our method, as implemented here, only permits vaccine descriptions where mechanisms are either absent, or present to the same extent, but does not accommodate more general situations where all vaccine mechanisms may be present with different intrinsic efficacies. Generalizing the approach here proposed to describe that situation should be equally feasible regardless of the distribution of protective vaccine effects among immunized individuals, especially for the analysis of balanced multi-centric trials [18,19].

The method proposed in this study can be used for interpreting RCTs for vaccine efficacy against TB conducted on IGRA+ individuals, and it can be extended to other trial designs, even for diseases obeying to different transmission dynamics structures. It can furthermore be used coupled with any transmission model of choice (see [17] for an exhaustive review of most recent modelling tools described in recent literature for TB), as long as it accommodates the description of the different routes to disease and mechanisms of action here described. By adapting our method to these situations, it will be possible to produce less arbitrary model-based impact forecasts based on all-or-nothing vaccine descriptions where the knowledge about the vaccine is incomplete.

## Materials and Methods

### Basal model calibration

The basic model describing TB acquisition of participants in the placebo arm during the trial (Figure 1A), can be expressed through the following system of ordinary differential equations:

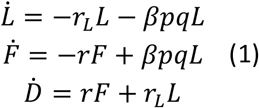

where we consider an endogenous LTBI reactivation rate centered around *r*_*L*_ = 7.5 · 10^−4^ *y*^−1^ (C.I. 6.37 · 10^−4^ 8.63 · 10^−4^) whereas fast progression rate to TB is centered in *r* = 0.9 *y*^−1^ (C.I. 0.765-1.035) [33]. According to [34], we consider that LTBI individuals have a 79% less risk of progressing to TB upon reinfection, that is, *q* = 0.21 (C.I. 0.14-0.30). Finally, the probability of fast progression is centered around *p* = 0.15 (C.I. 0.10-0.20) [21,35,36]. With these parameters fixed (drawn in each realization from normal distributions compatible with expected values and C.I.s), the transmission rate *β* is initially calibrated for each country, and within each age group, using a complete transmission model -the same used to evaluate vaccines impact (see sub-section on vaccine impacts below and supplementary information)-bound to fit the incidence and mortality burden reported by the WHO between 2000 and 2019 [23]. The same model is used to produce estimates of the fraction of fast vs. slow progressors among IGRA+ populations in each country and age group (see SI for the fitted values of *β* and the fractions of slow vs fast progressors per country and age group).

Using these dynamical parameters, the dynamics in the placebo arm of the trial are simulated, according to the system of ODEs in eq. (1), employing a version of the Gillespie algorithm where the reservoir F is mirrored in order to allow for independent tracking of the individuals undergoing primary TB who were initially in F as well as the individuals following the re-infection route to disease: L→F→D. As for individuals in the vaccine arms of the cohort, each all-or-nothing vaccine model can be mapped onto a combination of the epidemiological parameters (*r*_*L*_, *r*, or the product *βpq*, see supplementary table 2) being turned to zero for a fraction *ε* of the vaccinated individuals. Each in-silico trial comprehended the simulation of a fraction of individuals in different age groups, ranging from 18 to 50 years old, in three different African countries, with each country-age group combination being characterized by specific epidemiological parameters. We estimated these fractions from the reported participant distributions across age strata and country reported in [9] (See supplementary information for details). Once the result of each simulation is obtained through the Gillespie algorithm, the PoD efficacy is estimated as 1 minus the ratio of cases observed in the placebo and intervention arms.

We simulated N=10000 trials for each model and value of the intrinsic efficacy; with 200 values of the intrinsic efficacy ε uniformly distributed in the range [0,1]. Each one of these instances involves the simulation of the dynamics in both cohorts in three countries and within seven age-groups (16-20 to 46-50 years old, age-group width=5 years), that are combined into a single efficacy readout per instance. This yields a total number of trials simulated equal to 4.2 · 10^7^ per model, (200 intrinsic efficacies x 10.000 instances x 7 age groups x 3 countries); that is, 1.47 · 10^8^ trials simulated in total for the 7 models analyzed.

### Bayesian analyses

Let us consider the likelihood *P*(*VE*_*dis*_= 49.7%|*i, ε*) that each of the possible models defined by the combination of parameters {*i, ε*}, (where the integer index *i* ∈ {1,2, …, 7} determines the specific vaccine mechanism(s) at work (i.e. the vaccine model type), and the continuous parameter *ε* ∈ [0,1] captures its intrinsic efficacy) generates a PoD efficacy estimate compatible with the one observed for M72/AS01_E_. Using this likelihood term, we apply Bayes rule to define the posterior probability associated to each particular model:

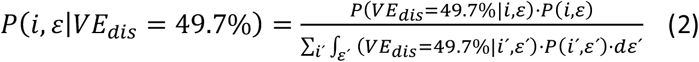

And derive a model-type posterior probability, by integrating over all possible intrinsic efficacy values as follows:

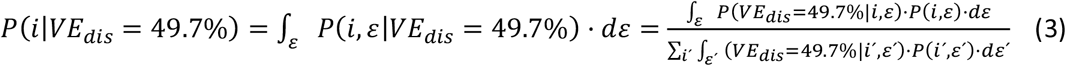

If we consider uniform non-informative priors in eq. (3) (that is *P*(*i, ε*) = *P*(*i*′, *ε*′) ∀ (*i, i*^′^, *ε, ε*^′^)), the model-type posterior can be obtained from the marginalized likelihoods:

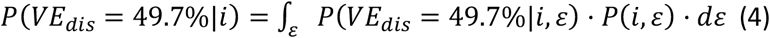

Which we estimate from the density distributions of the PoD efficacy readouts *VE*_*dis*_obtained from each model using the value of Kernel density estimators (R package KerSmooth) of the frequency of trials evaluated at *VE*_*dis*_= 49.7%. Plugging the numerical estimates of *P*(*VE*_*dis*_= 49.7%|*i*) into eq. (3) allow us to quantify the relative support in the data for the seven different vaccine descriptions provided. Confidence intervals for these model posterior estimates represented in figure 3b are obtained by bootstrapping the calculations N=5000 times, each of which is obtained by sampling with replacement N=10.000 trial simulations.

Then, we also estimate the intrinsic efficacy values *ε* that are most compatible with the given observed efficacy-against-disease readout *VE*_*dis*_= 49.7% under each of the model type descriptions, this time applying the Bayes rule over each model type independently:

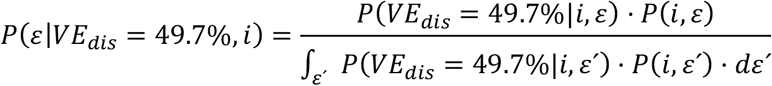

Where likelihood terms *P*(*VE*_*dis*_= 49.7%|*i, ε*) are estimated from the simulations using Kernel density estimates obtained for each of the N=200 values of *ε* covered. The first momentum of these posterior distributions corresponds to the expected values of the intrinsic efficacy parameter under each model type, that is:

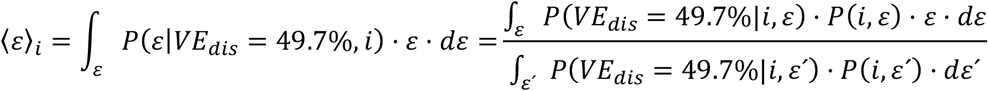

which is captured in figure 3C, along with its confidence intervals obtained from the posterior distribution *P*(*ε*|*VE*_*dis*_= 49.7%, *i*) itself, fitted to a normal distribution.

Finally, we build model-based Bayesian estimates of vaccine impact as a weighted linear combination of the impacts foreseen by each type of vaccine, expressed as incidence reduction rates, as follows:

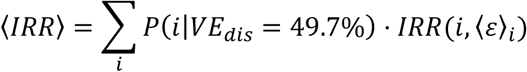

where the averaged incidence reduction rates *IRR*(*i*, ⟨*ε*⟩_*i*_) are computed using the same comprehensive transmission model used to estimate transmission rates *β* and fractions of prevalent fast vs slow progressors [23].

### Impact evaluations

In this study, we make use of the transmission model introduced in [23], and generalized to describe the introduction of novel vaccines in [16]. This model constitutes a conceptual extension of the basic model sketched in figure 1A, where different types of disease are considered, along with treatment outcome dynamics and eventual relapses. The model integrates demographic data and empiric mixing patterns among age strata along with epidemiological parameters and TB burden trends (incidence and mortality), in order to produce baseline incidence and mortality forecasts per country, as well as vaccine impact evaluations. For further details on the model the reader is referred to the supplementary information appendix, along with references [16,23].

## Data Availability

Code with the implementation of the novel methods introduced in this study is available at https://github.com/MarioTovarCalonge/IGRA_positive_analysis.

## Acknowledgements

We thank M. Gutiérrez for graphic design assistance.

## Funding

Government of Aragón PhD contract order IIU/796/2019 (MT), grant E36-20R (FENOL: MT, YM, JS). MCIN/AEI/ 10.13039/501100011033 and “ESF Investing in your future”: grant PID2020-115800GB-I00 (YM), and grants PID2019-106859GA-I00 and RYC-2017-23560 (JS).

Banco Santander: Santander-UZ 2020/0274 (YM)

Soremartec S.A. and Soremartec Italia, Ferrero Group. (YM)

The funders had no role in study design, data collection, and analysis, decision to publish, or preparation of the manuscript.

## Author Contributions

MT, JS and YM designed research; MT performed research; MT and JS analyzed data, and discussed results with YM. JS and MT wrote the article with inputs from YM. All authors read and approved the final version of the manuscript.

## Data and materials availability

Code with the implementation of the novel methods introduced in this study is available at https://github.com/MarioTovarCalonge/IGRA_positive_analysis. Those include algorithms written in R (tested in version 3.6.3 2022-05-25) and C language, with dependences to the following specific libraries: fanplot, ggplot2, gridExtra, kdensity, KernSmooth, from R. Fanplot, ggplot and gridExtra are used for visualization purposes. kDensity and KernSmooth are used for the Bayesian analysis.

## Supplementary Materials

Supplementary_Text_S1.pdf

