## Supplementary_Text_S1 for "Identifying vaccine-mechanism bias in mathematical models of vaccine impact: the case of tuberculosis"

Mario Tovar,[1,2] Yamir Moreno,[1,2,3] and Joaquín Sanz[1,2]

[1] *Institute for Biocomputation and Physics of Complex Systems (BIFI), University of Zaragoza, Zaragoza 50009, Spain*

[2] *Department of Theoretical Physics, University of Zaragoza, Zaragoza 50009, Spain*

[3] *ISI Foundation, Via Chisola 5, 10126 Torino, Italy*

**Contents**

I. Methods 2
A. In silico clinical trial simulations: Placebo arms 2
B. In silico clinical trial simulations: Vaccine arms 3
C. Model-based impact evaluations of TB vaccines 5
1. *M.tb.* transmission dynamics within age strata. 5
2. Population dynamics across age strata: ageing and demographic evolution. 6
3. Vaccine descriptions 7
4. Uncertainty estimates for model-based estimates 7

References 10

**List of Figures**

1 Percentage of fast progressors in Kenya, South Africa and Zambia, estimated in 2015. . . . . . . . . . . . . . 4
2 Natural History schemes of the TB spreading model . . . . . . . . . . . . . . . . . . . . . . . . . . . . . . . . . . . . . . . . . . . . 8

**List of Tables**

I Parameters used in Gillespie algorithm. . . . . . . . . . . . . . . . . . . . . . . . . . . . . . . . . . . . . . . . . . . . . . . . . . . . . 3
II Calibrated values of force of infection per age and per country. . . . . . . . . . . . . . . . . . . . . . . . . . . . . . . . . . 3

### I. Methods

#### A. In silico clinical trial simulations: Placebo arms

The disease dynamics of the control arm in a randomized clinical trial for vaccine efficacy against tuberculosis (TB) conducted on IGRA-positive individuals is described by the following system of ordinary differential equations (ODEs):

$$\dot{L} = -r_L L - \beta p q L \tag{1}$$

$$\dot{F} = -rF + \beta p q L \tag{2}$$

$$\dot{D} = rF + r_L L \tag{3}$$

where we have three states (fast latency $F$, slow latency $L$ and active disease $D$), five epidemiological parameters (infection rate $\beta$, fast (slow) progression to disease rates $r$ ($r_L$), probability of fast progression upon infection $p$, and risk reduction for fast progression upon re-infection $q$), and three different types of transitions between them (events): reinfections ($L- > F$), slow progression to disease ($L- > D$) and fast progression to disease ($F- > D$). The sequence of events of each type during the follow up of the study is modelled stochastically, using an implementation of the Gillespie algorithm where the daily probabilities (propensities) of each type of transition are: $a_{L\to F} = \beta pqL$, $a_{L\to D} = r_L \cdot L$, and $a_F = r \cdot F$, respectively. In our implementation, the reservoir $F$ is duplicated: one instance contains only the individuals already in $F$ at the beginning of the study, while the second one starts empty, and receives the eventual cases of reinfections that will undergo fast progression. In this way we can keep track of all three different routes to disease independently without altering the dynamics.

The algorithm works, by iterating, at each time step $t$, the following operations:

1. Calculate the probabilities $a_j$ of each type of event happening at $t$.
2. Generate an exponentially distributed random variable $dt = -\frac{log(r_n)}{R}$, where $r_n$ is a random number uniformly distributed in the interval $(0, 1)$ and $R$ is the sum of the probabilities of all events at time $t$. The next event will occur at $t' = t + dt$.
3. Determine the event to occur by stochastically drawing a process from all possible processes according to their respective probabilities $a_j$.
4. Update the population according to the event that has taken place.
   - If a reinfection takes place, then $L(t + dt) = L(t) - 1$ and $F(t + dt) = F(t) + 1$
   - If an endogenous reactivation occurs, then $L(t + dt) = L(t) - 1$ and $D(t + dt) = D(t) + 1$.
   - If a fast transition to disease happens, then $F(t + dt) = F(t) - 1$ and $D(t + dt) = D(t) + 1$.
5. Move to the next time step, $t = t + dt$.

To implement this algorithm, we need two ingredients, namely: the epidemiological parameters ($r$, $r_L$, $p$, $q$, and $\beta$), and the initial conditions of the system $L(t = 0)$ and $F(t = 0)$ ($D(t = 0) = 0$ since individuals with signs of active TB are excluded from the type of trial we analyze).

All epidemiological parameters but the infection rates are assumed to have the same values in all countries and age groups analyzed, and are taken from the literature, as reported in Supplementary Table I. In order to estimate the force of infection $\beta$ in each country and age group, we capitalize on the comprehensive spreading model described in reference [6], which we calibrate in each country appearing in the study, (South Africa, Kenya and Zambia) to record the calibrated force of infection, averaged during 2015. This procedure is done per age group, and per country, and the resulting values are used later in the Gillespie algorithm. Calibrated rates for $\beta$ are shown in Supplementary Table II.

Finally, we also used the model to obtain estimates of the relative fraction of latent individuals without a past of active TB that show a high risk of progressing to active TB in the next 12-24 months (fast progressors) and those for whom that risk is much lower (slow progressors). The results of this exercise, for each country and age-group, are represented in figure 1.

| Parameter | Value | Reference |
|---|---|---|
| $p$ | 0.150 (0.100-0.200) | [3–6] |
| $q$ | 0.210 (0.140-0.300) | [6, 7] |
| $r$ | 0.900 (0.765-1.035) | [2, 6] |
| $r_l$ | $7.5 \cdot 10^{-4}$ ($6.4 \cdot 10^{-4}$-$8.6 \cdot 10^{-4}$) | [2, 6] |

*Table I: Parameters used in Gillespie algorithm.*

| age group | Kenya | Zambia | South Africa |
|---|---|---|---|
| 0 | 0.018 | 0.061 | 0.060 |
| 1 | 0.031 | 0.129 | 0.101 |
| 2 | 0.039 | 0.159 | 0.126 |
| 3 | 0.046 | 0.139 | 0.150 |
| 4 | 0.038 | 0.097 | 0.134 |
| 5 | 0.033 | 0.082 | 0.118 |
| 6 | 0.032 | 0.083 | 0.117 |
| 7 | 0.038 | 0.095 | 0.137 |
| 8 | 0.039 | 0.098 | 0.144 |
| 9 | 0.043 | 0.105 | 0.160 |
| 10 | 0.044 | 0.109 | 0.164 |
| 11 | 0.041 | 0.102 | 0.153 |
| 12 | 0.045 | 0.112 | 0.170 |
| 13 | 0.049 | 0.125 | 0.180 |
| 14 | 0.038 | 0.098 | 0.140 |

*Table II: Calibrated values of force of infection per age and per country.*

##### B. In silico clinical trial simulations: Vaccine arms

The previous section describes the algorithmic strategy used to simulate the evolution of the placebo arms of the simulated trials. In order to simulate the vaccine arm, we considered vaccines featuring three basic mechanisms of action, either alone, or combined (7 models). These mechanisms correspond to the ability of the vaccine to effectively block, independently, each of the transition types ($L->F->D$, $L->D$, or $F->D$) in a fraction equal to $\varepsilon$ of all vaccinated individuals, according to an all-or-nothing vaccine description. Therefore, in each of the models, a parallel compartmental model can be derived, not for all the vaccinated individuals, but only for the fraction $\varepsilon$ of them who is effectively protected.

In what follows we describe all these compartmental models, and how are are we using them to integrate the description of the dynamics of the protected individuals within our simulation framework.

*a. Model 1* In this vaccine model, the transition $F->D$ of individuals who already were in $F$ at the beginning of the study is halted. This is modeled by shifting the protected individuals from $F$ to $L$ right after their vaccination, therefore assuming that vaccine protection implies that the risk of fast progression is substituted with the much lower risk of slow progression after endogenous reactivation. This way, the dynamical rules for the time evolution of this arm remain unchanged with respect to those described above for the placebo arm, and the difference between arms comes from the fact that the fast progression reservoir is emptied right after vaccination in the intervention arm.

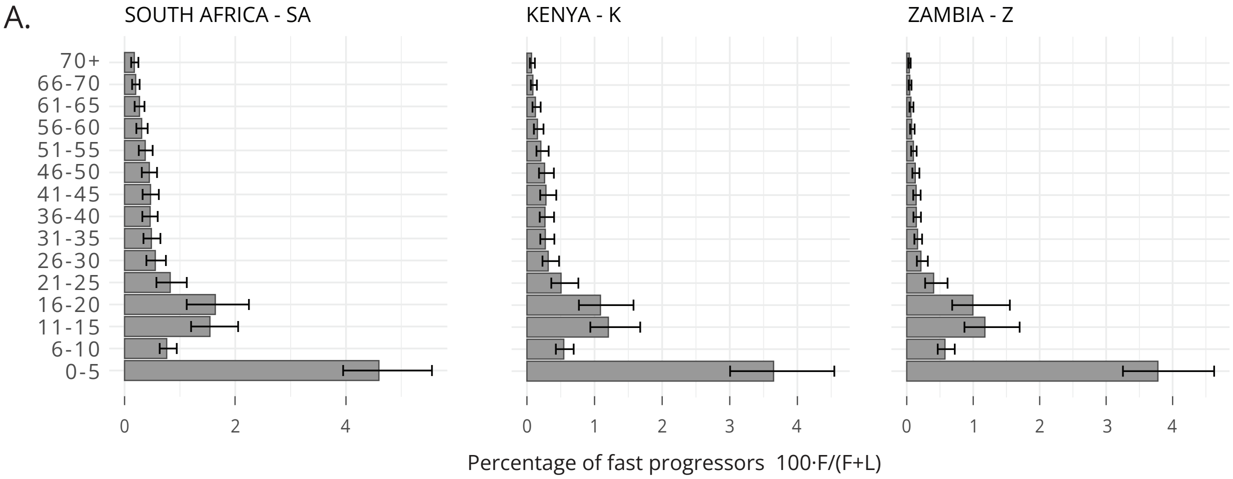


*Figure 1: Percentage of fast progressors in Kenya, South Africa and Zambia, estimated in 2015.*

*b. Model 2* In this model the vaccine is able to completely stop the reinfections from **L** to **F**, which is described by the subsequent ODEs for the $\varepsilon$ fraction of vaccinated individuals:

$$\dot{L} = -r_L L \tag{4}$$
$$\dot{F} = -rF \tag{5}$$
$$\dot{D} = rF + r_L L \tag{6}$$

The $\varepsilon$ fraction of individuals that is protected by the vaccine is simulated using a variant of the Gillespie algorithm where the reinfection event is not considered. Now, each time that a new time is selected by the algorithm, the event that takes place is selected only between endogenous reactivation and fast progression.

*c. Model 3* In this model the vaccine has the ability to interrupt the endogenous reactivation process, thus preventing **L** individuals to progress to disease. This situation is described with ODEs for the $\varepsilon$ fraction of vaccinated individuals that is protected, as follows:

$$\dot{L} = \beta pqL \tag{7}$$
$$\dot{F} = -rF + \beta pqL \tag{8}$$
$$\dot{D} = rF \tag{9}$$

Here, each time that a new time is selected by the Gillespie algorithm, the event that takes place is selected only between reinfection and fast progression.

*d. Model 4* Here we assume that the vaccine is able to completely stop the reinfections from **L** to **F**, which is described by the same ODEs of second model, as well as another effect that mimics model 1. We have then a shift in the initial distribution of participants from **F** to **L** in the protection cohort, combined with a perfect protection versus reinfections.

*e. Model 5* Here we consider that the vaccine is able to completely stop the endogenous reactivations, which is described by the same ODEs of the third model, and also has the effect of model 1. We have then a shift in the initial distribution of participants from **F** to **L** in the protection cohort combined with a perfect protection against disease for **L** individuals.

*f. Model 6* This model combines the effect of the models 2 and 3, acting at the same time over endogenous and exogenous reactivation. This situation is described with ODE's as follows:

$$\dot{L} = 0 \tag{10}$$

$$\dot{F} = -rF \tag{11}$$

$$\dot{D} = rF \tag{12}$$

The Gillespie algorithm here is the most simple one we can have, as only an event can take place, so each new time that an event occur, a fast progression event happens.

*g. Model 7* Finally, in this model we are considering that the vaccine holds a combination of all the three basic effects depicted in models 1 to 3, thus being the most powerful vaccine among all the models, for the same intrinsic efficacy $\varepsilon$. The dynamical behaviour is the same that we have in model 6 combined with the fact that several **F** individuals are moved towards **L** at the start of the trial.

Following the algorithms specified above to model the dynamics in the placebo and vaccine arms, informed with the initial conditions and epidemiological parameters as described in the previous section, we implement a number of $N = 2 \cdot 10^6$ independent simulations for each country and age-group combination, distributed along the entire range of possible intrinsic vaccine efficacies $\varepsilon \in [0, 1]$. Once the Gillespie algorithm is implemented in the three countries and all age strata from $16 - 20$ to $46 - 50$, all results are averaged, weighted by the relative representation of individuals from each country and age strata in the $M72/AS01_E$ trial, to obtain a $\mathrm{VE}_{dis}$ outcome from each simulation. In each simulated instance, it is important to highlight that all the parameters and initial conditions are stochastically draw from their corresponding distributions, given their central values and confidence intervals.

##### C. Model-based impact evaluations of TB vaccines

In this study we capitalized on the work described in [6] to adapt the model proposed there in order to estimate impacts of different vaccines, in terms of their ability to reduce the incidence rate upon their introduction in nation-scale settings. The adapted version introduces an improved strategy for the assessment of outcomes' confidence intervals and their sensitivity to inputs uncertainty.

The model is a deterministic, age-structured tool based on ordinary differential equations, where individuals belonging to different age-strata are considered to experiment different levels of epidemiological risk that translate into age-specific parameter values for some of the key dynamical processes. Social mixing patterns driving the contagion dynamics are modelled using empiric survey data, and an explicit coupling between populations' evolution and transmission dynamics allows an explicit description of the effects of demographic ageing (past and foreseen), and the evolution of the epidemics.

Using this model, we performed two tasks. First, the model was calibrated in the countries where the $M72/AS01_E$ trial took place: South Africa, Kenya and Zambia, as described above, in order to obtain estimates of infection rates per age group and country in 2015, as well as fractions of LTBI prevalence in that year corresponding to the fast vs slow latent TB reservoirs. These estimates were used to feed the trial simulations from which the seven vaccine descriptions described in the main text were achieved. Second, the model was used to simulate the introduction of those vaccines in a different set of countries: India, Indonesia and Ethiopia,. The vaccination campaigns simulated starts at the end of 2025, and the target populations are adolescents between 15-20 years old. Using these simulations, we evaluated the impacts of each vaccine as the incidence rate reduction achieved by 2050.

In the following lines we summarize the main aspects of the model; for further detail, the reader is referred to [1, 6].

###### *1.* M.tb. *transmission dynamics within age strata.*

The model includes 15 different age groups, 14 of them covering 5 years of age up to 70 years old, and the last one containing all individuals older than 70 years old. Within each age group we consider two different branches of individuals: protected, and non-protected by the vaccine.

Within each of these branches we have a state for unexposed individuals –susceptible–, that can get infected and progress through two different latency paths to disease –fast and slow –. Once in disease, we distinguish different kinds of disease, depending on its aetiology: -non pulmonary, pulmonary (smear positive) and pulmonary (smear negative)–. Then the individuals can get diagnosed at a calibrated rate and depending on whether they are left untreated or treated, remain in disease or progress to the treatment state, respectively. Right after all the disease phase we consider separately the treatment outcomes contemplated by the WHO data schemes: treatment completion, default, failure and death [11, 12].

As a summary, there are several types of possible transitions between the states described above and included in figure 2**A**:

- Infection processes: after a contact with an infectious individual, susceptible individuals ($S$) can get infected, entering either the fast ($L_f$), or slow latency states ($L_s$).
- Re-infection processes: individuals in the slow latency reservoir can get re-infected, a fraction of which will develop TB fast after re-infection. This is modelled here as a transition from $L_s$ to $L_f$.
- Development of active TB: infected individuals (those in $L_s$ or $L_f$) might develop initially undiagnosed -and thus untreated- TB (progressing to $D$).
- TB diagnosis: with some delay after the disease onset, TB gets diagnosed and treatment starts (transition $D$ to $T$)
- Spontaneous recovery: (transitions from $D$ to $R$)
- Treatment outcomes: (transitions from $T$ to $R$) different possible outcomes are possible: –either success or failure/default-
- Disease relapse: (transitions back from $R$ to $D$)
- Death: active TB patients, either diagnosed or not, are assigned with a TB specific mortality rate.

Back to the dynamical description, this scheme defines the time evolution of individuals inside a given age strata, according to a set of differential equations that are exhaustively disclosed in [6].

Within this scheme, infections may occur after a contact between susceptible individuals and infectious ones. Let $S(a,t)$ represents the number of susceptible subjects in age group $a$, at a given time $t$, the number of new infections that will be observed will be equal to the product of $S(a,t)$ and the force of infection perceived by that sub-population, $\lambda(a,t)$, which represents the fraction of susceptible individuals who get infected per year. The force of infection is proportional to the following weighted sum:

$$\sum_{a'} \xi_c(a,a',t)\Upsilon(a',t) \tag{13}$$

where $\Upsilon(a',t)$ is the density of all the infectious individuals within age-group $a'$ at time step $t$, weighted by their relative infectiousness, and $\xi_c(a,a',t)$ represents the relative contact frequency that an individual of age $a$ has with individuals of age $a'$ at time $t$, with respect to the overall average of contacts that an individual has per unit time with anyone else, as described in [6].

For the computation of the contact matrices used in our model, we have integrated data from different survey studies conducted in several countries across the world for building contact matrices that we can use distinguishing world regions. Our description of the age-dependent contact patterns driving contagion takes into account that, as the demographic structure of the population changes, the contact patterns change too [19], implying that our matrices are time-evolving, as described in [6].

##### *2. Population dynamics across age strata: ageing and demographic evolution.*

This model performs the simultaneous description of the disease dynamics across all age-groups in an entire population, for which it uses parameters that are, in general, dependent on age. Therefore, it is not enough to describe how the sub-populations associated to the disease states evolve in time as individuals age to properly describe the evolution of the individuals the model also includes ageing dynamics whereby individuals transit across the different age-strata as they get older, which introduces a driver of demographic evolution in the population under study.

In order to render that evolution compatible with demographic dynamics observed in the populations under analysis, we use demographic data and prospects from the UN population division [13]. More specifically, at each time step, we introduce continuous correction terms $\Delta_N(a,t)$ that are added or subtracted from the population within the age stratum $a$ at time $t$ while the simulation unfolds. These terms are calculated dynamically to make the time evolution of the demographic pyramid match the demographic forecasts reported in the United Nations population division database until the end of the simulation [13]. Proceeding this way, the correction terms $\Delta_N(a,t)$ are distributed among all disease states proportionally to their relative size. Note that these terms represent changes in the population of each strata that are unrelated to the dynamics of the disease (TB unrelated mortality and migratory fluxes).

##### *3. Vaccine descriptions*

Vaccine-mediated protection is described, within the framework of our model, in a parallel cohort where the dynamics of protected individuals unfold (see figure 2 **B**). Here we study seven different vaccines, whose mechanisms of action are described above. Protection induced by these vaccines is described according to an all-or-nothing scheme, meaning that an $\varepsilon$ fraction of the vaccinated individuals is fully protected against the routes to disease blocked by the vaccine (and only against them), while the rest remains susceptible. For the sake of our analyses, we have assumed that the protection happens irrespective of whether individuals were infected or not before vaccination in each case.

The architecture of the possible transitions between disease related states in each of the model is identical, with the exception of the fate reservoir of individuals who were fast progressors at the moment of vaccinations. In the models where the vaccine is assumed to protect against on-going fast progression to disease, linked to a exposure that happened short before vaccination (models 1,4,5, and 7), vaccinated individuals in $L_f$ are shifted to the reservoir of slow protected progressors $L_s^v$ upon vaccination (see figure 2 **A**), while, for the rest of models (models 2,3,6), they are moved to $L_f^v$.

In what regards the other two mechanisms in the vaccine models (arresting of fast progression upon infection, and arrest of endogenous reactivation), their inclusion in the models is done by setting specific parameters to zero, effectively blocking the transitions controlled by them without the need of modifying model archictecture. Vaccine models including the arrest of slow progression to disease (models 3,5,6,7) see the corresponding rate $r_L$ set to zero, while vaccine models including protection against reinfection are parametrized by assuming no re-infection risk, that is $q = 0$ (models 2,4,6,7).

In each case,, a vaccination campaign is considered, focused on adolescents between 15-20 years old, that are targetted at once in 2025, at the same time that new individuals entering this age group as time passes by are progressively, and uninterruptedly vaccinated until 2050. Assuming a perfect coverage, within an all-or-nothing conception, the intrinsic efficacy $\varepsilon$ defines the fraction of vaccinated individuals that gain protection, transiting to the protected branch of the model.

##### *4. Uncertainty estimates for model-based estimates*

Uncertainty in the outcomes from our model come from the following independent sources:

- Uncertainty in parameters associated with the Natural History of the disease.
- Uncertainties in TB burden estimations provided by the WHO. Based upon a number of case notifications surveilled in each country, the World Health Organization provides estimations for incidence and mortality rates for the countries under analysis, which feature significant uncertainty bands which we propagate to our model outcomes.
- Demographic structures, which are considered as a single uncertainty source. Those demographic structures are reported in the form of demographic pyramids with 15 age groups, first 14 from 0y to 70y each 5y and the last one comprises 70+y. Each group has its own uncertainty, but said uncertainty levels are not independent among them, which we take into account in our analyses.
- Contact matrices, whose uncertainty comes from the variability between studies used for building aggregated matrices, as described in [6].

Considered these independent uncertainty sources, we propagate them to our model outcomes by generating a set of possible modelling runs where each escalar (i.e. one-dimensional) parameter is independently sampled from a distribution informed by its confidence intervals.

In what regards multi-dimensional inputs -that is, TB incidence and mortality series from WHO data, demographic pyramids and contact matrices, the eventual co-dependence of their elements is treated differently in each case.

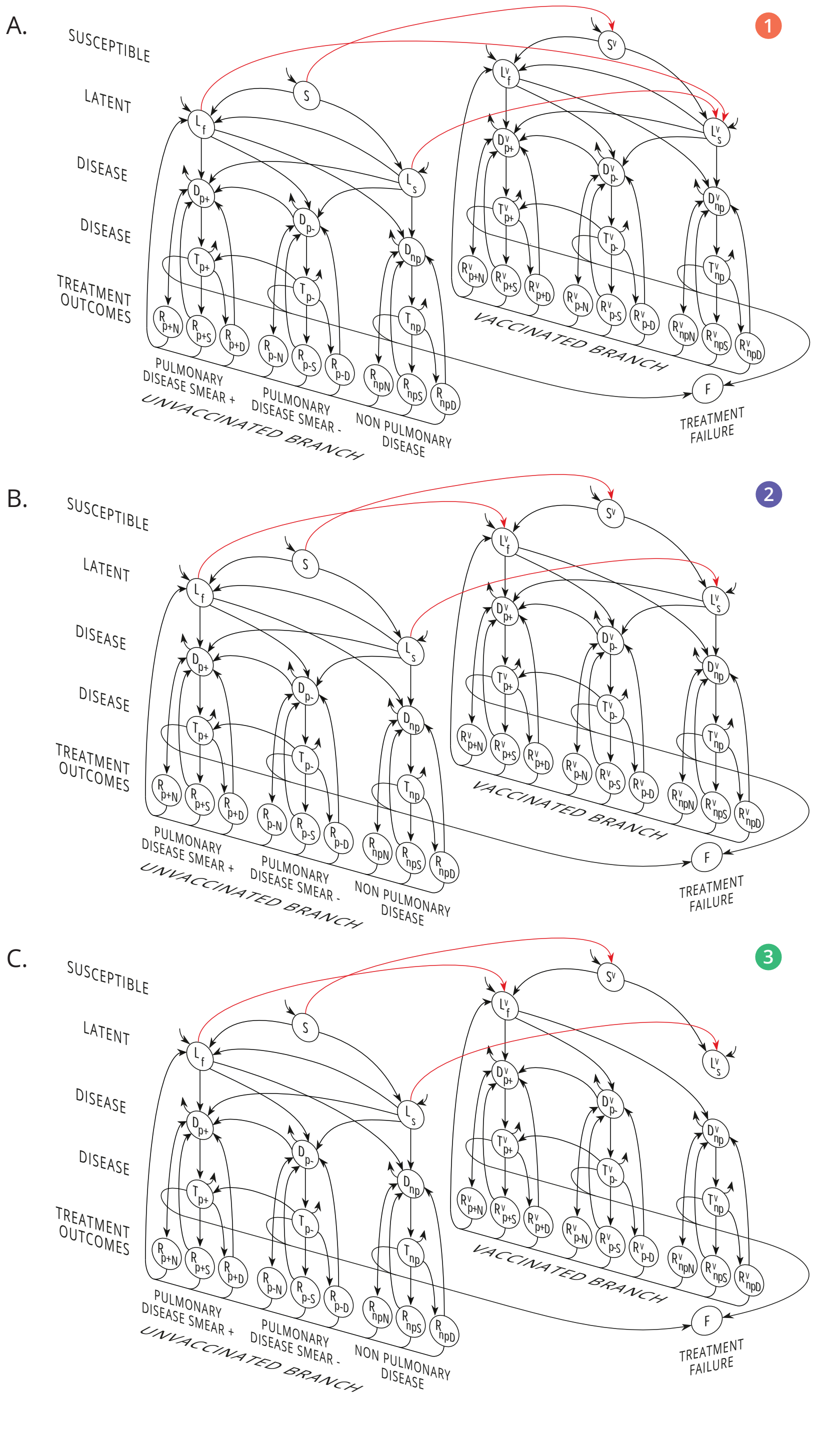


*Figure 2: Natural History scheme of the TB spreading model. S: susceptible. L: latent. D: (untreated) disease, T (treated) disease, R recovered, F: failed recovery. Types of TB considered: p+: Pulmonary Smear-Positive, p-: Pulmonary Smear-Negative, np: Non-pulmonary. Treatment outcomes: RN: Natural recovery, RS: Successful treatment, RD: Default (abandon of treatment), F: treatment failure. Two branches are modelled: the placebo arm (i.e. non-vaccinated, or vaccinated but non-protected), and a parallel branch where individuals are protected by the effect of the vaccine. Vaccination transitions between cohorts are therefore included (i.e. red arrow transitions), and each panel features a vaccine arm corresponding to the first three models contemplated in the article, describing vaccines conferring protection against only one of the three routes to disease: (A) Model structure 1: the vaccine protects against primary TB upon first infection. This is implemented -analogously to what is done in the Gillespie model used to simulate the trials- by shifting individuals in the reservoir* $L_F$ *to the reservoir* $L_S^v$ *as individuals enter into the target age-group* $(15-20$ *years old) of the vaccination campaign. (B) Model structure 2: the vaccine protects against re-infection, which is implemented by blocking the transition* $L_S^v$ *to* $L_F^v$ *for all age strata from the target group onward (C) Model structure 3: the vaccine protects against endogenous reactivation of LTBI, achieved by blocking the direct transitions from* $L_S^v$ *to the active disease reservoirs* $D^v$ *for all age strata from the target group onward. Model structures 4 to 7 can be trivially obtained by combining these three.*

Uncertainty in contact matrices is estimated from the heterogeneity in the reported results of the different studies used to obtain the aggregated matrices used here. In the case of African countries we combine surveys in Kenya [14], Zimbabwe [15] and Uganda [16] to obtain a unique matrix broadly representative of contact structures in Africa, as described in [6, 19], while for countries in Asia we use surveys conducted in China [17] and in Japan [18] that are combined for getting a matrix usable in the Asian region. In this case, the confidence intervals of each field in the matrices is obtained, and matrices are re-sampled according to them, assuming that matrix fields (corresponding to the results of the surveys on different sub-populations) are mutually independent.

Second, in order to capture the co-dependence of the methods used by the WHO to produce the estiomates of TB incidence and mortality rates that we use in our model forecasts, we re-sample alternative burden scenarios by drawing numbers from an standard normal distribution, which are later scaled to generate the corresponding incidence $i$ of mortality $m$ series, in each realization, as follows:

$$i^j_{\text{new}} = i^j_{\text{orig}} + Z \cdot \sigma^j_i \tag{14}$$

$$m^j_{\text{new}} = m^j_{\text{orig}} + Z \cdot \sigma^j_m \tag{15}$$

where $\sigma^j_i$ and $\sigma^j_m$ are the standard deviations of the incidence and mortality levels, respectively, that are associated with the 95% confidence intervals reported by the WHO at the j-th year.

Finally, in order to model uncertainty coming from demographic data, we decided to sample alternative demographic pyramids where random, normally distributed noise is considered in the proportion of younger vs older individuals, distributed across age strata in a linear way. To do so, in each iteration we draw a single Z score from a standard normal distribution, and generate a new base of the demographic pyramid, $n(a=1)_{\text{new}}$, as follows:

$$n(a=1)_{\text{new}} = n(a=1)_{\text{orig}} + Z \cdot \sigma(a=1) \tag{16}$$

Once this is done, we modify the rest of the age groups from the base to the top of the pyramid using age-dependent Z-scores $Z(a)$ that are re-scaled linearly to ensure that, while $Z(1) = Z$, the final $Z(15) = -Z$. This is done linearly, ensuring that the individuals added, or removed, at the younger strata are compensated by statistically analogous deviation, in the opposite direction, in the older strata, by applying the following rule:

$$Z(a, Z') = M(Z') \cdot a + K(Z') \tag{17}$$

where $M(Z') = -\frac{1}{7}Z'$ and $K(Z') = \frac{8}{7}Z'$.

Consequently, the new value for any age-group is given by:

$$n(a)_{\text{new}} = n(a)_{\text{orig}} + Z(a, Z') \cdot \sigma(a) \tag{18}$$

This sampling procedure is repeated for all parameters and sources of uncertainty a number $N = 500$ times, after each of which, the model is re-callibrated to generate an ensemble of possible baseline trend simulations.

Then, on each of the instances of this ensemble, we introduce vaccines according to the $n = 7$ different vaccine types considered in the study, and estimate their respective impact, measured as the foreseen incidence rate reduction at 2050. If we are considering vaccine type $i \in [1, 7]$, and model-run $v \in [1, 500]$, we will denote the corresponding impact as $IRR(i, \langle\epsilon\rangle_i, v)$, where $\langle\epsilon\rangle_i$ is the expected value of the efficacy parameter for model $i$, as represented in figure 3C.

In turn, in a final step, we combine the results of all models in a weighted sum performed for each realization that uses bayesian posteriors as weights to obtain an overall estimate of impact $\langle IRR(v)\rangle$, with $v \in [1, 500]$, that is, in each case, agnostic to the vaccine mechanism. These Bayesian averages are obtained as follows:

$$\langle IRR(v)\rangle = \frac{\sum_{i=1}^{7} I(i,v)P(i,v|VE_{\mathrm{dis}} = 49.7\%)}{\sum_{i=1}^{7} P(i,v|VE_{\mathrm{dis}} = 49.7\%)} \tag{19}$$

Where the posteriors $P(i,v|VE_{\mathrm{dis}} = 49.7\%)$ used in each case are drawn stochastically from the marginal posterior distributions represented in figure 3B in the main text, independently for each realization $v \in [1, 500]$.

Finally, median and confidence intervals for the impacts derived from each particular model, as well as the averaged impacts are extracted from the vectors of $N = 500$ stochastic estimates for each of them. When comparing impacts derived from two vaccine models, it is important to highlight that the differences are paired, in such a way that the differences in impact between two given vaccine models are evaluated $N = 500$ times, in each of which both vaccines are compared against the same baseline trend.

---

[12] World Health Organization Global tuberculosis report 2019, 2019, Geneva

[13] Population Division Database, http://esa.un.org/unpd/wpp/index.htm (accessed November 2016), 2016

[14] Kiti, Moses Chapa and Kinyanjui, Timothy Muiruri and Koech, Dorothy Chelagat and Munywoki, Patrick Kiio and Medley, Graham Francis and Nokes, David James, Quantifying age-related rates of social contact using diaries in a rural coastal population of Kenya, PloS one, 9(8), e104786, 2014, Public Library of Science

[15] Melegaro, Alessia and Del Fava, Emanuele and Poletti, Piero and Merler, Stefano and Nyamukapa, Constance and Williams, John and Gregson, Simon and Manfredi, Piero Social Contact Structures and Time Use Patterns in the Manicaland Province of Zimbabwe, PloS one, 12 (1), e0170459, 2017, Public Library of Science

[16] le Polain de Waroux, Olivier and Cohuet, Sandra and Ndazima, Donny and Kucharski, Adam and Juan-Giner, Aitana and Flasche, Stefan and Tumwesigye, Elioda and Arinaitwe, Rinah and Mwanga-Amumpaire, Juliet and Boum, Yap and others Characteristics Of Human Encounters And Social Mixing Patterns Relevant To Infectious Diseases Spread By Close Contact: A Survey In Southwest Uganda, bioRxiv, 121665, 2017, Cold Spring Harbor Labs Journals

[17] Read Jonathan M., Lessler Justin, Riley Steven, Wang Shuying, Tan Li Jiu, Kwok Kin On, Guan Yi, Jiang Chao Qiang and Cummings Derek A. T. Social mixing patterns in rural and urban areas of southern China , Proc. R. Soc. B.28120140268 http://doi.org/10.1098/rspb.2014.0268

[18] Ibuka Y, Ohkusa Y, Sugawara T, et al Social contacts, vaccination decisions and influenza in Japan, J Epidemiol Community Health 2016;70:162-167.

[19] Arregui, S, Aleta, A, Sanz, J, and Moreno, Y. Projecting social contact matrices to different demographicstructures,PLoS Comput Biol, 14(12), e1006638, 10.1371/journal.pcbi.1006638, (2018)
